# Knowledge, Perception, and Preventive Practices of Livestock Workers and Household Animal Owners Regarding Anthrax in Nigeria

**DOI:** 10.1101/2024.02.26.24303419

**Authors:** E. Cadmus, H.K. Adesokan, E.J. Awosanya, P.M. Iziomo, V.O. Akinseye, M.A. Besong, A.O. Jolaoso, Nma Bida A., J.P. Akangbou, E. Nwanga, G T. Orum, A.O Omileye, A.A. Adeleye, S. Owoicho, O.J. Taiwo, R. Ansumana, C. Vakuru, S.I. Cadmus

## Abstract

Anthrax disease outbreak is a significant public health and socioeconomic problem, especially in low and middle-income countries (LMIC) like Nigeria. Inadequate knowledge and poor preventive practices against the disease among livestock workers and household animal owners remain important for disease transmission. Following the recent outbreaks in Nigeria, a cross-sectional study was carried out to assess the knowledge, perception and preventive practices of livestock workers and household animal owners regarding anthrax and the associated socioeconomic implications in Nigeria.

A pretested, semi-structured, interviewer-administered questionnaire was used to elicit relevant information from the respondents (n=1025) in seven of the 36 states in Nigeria. Data were analysed using SPSS version 29. Univariate analysis was done and Chi-square test statistics was test for association between the knowledge/perception and other variables.

Of the 1025 respondents, 58.6% and 79.9% demonstrated good knowledge and positive perception towards anthrax. However, there were important exposure practices, including a lack of preventive measures against anthrax infection (22.0%). Besides, only 27.7% of the respondents knew about the anthrax vaccination programme for livestock in the study area. With respect to the socioeconomic effects of the disease outbreak, 23.8% of the respondents indicated that the regulations imposed during an anthrax outbreak affect their livestock-related activities, while 40.6% were worried they might go out of business due to the anthrax outbreak. The respondents’ knowledge of anthrax was significantly associated with higher education (p=0.000), level of awareness (p=0.000) and perception of risk (p=0.000).

The study reveals a relatively high level of perception but an average knowledge level regarding anthrax with associated socioeconomic impacts among livestock workers and household animal owners in Nigeria. An important knowledge gap includes the poor knowledge of the routine annual vaccination of animals. Hence, mitigation strategies should include educational programmes targeting this gap.

## Introduction

Anthrax is a zoonotic disease of significant public health importance caused by a gram-positive, rod-shaped bacterium, *Bacillus anthracis* [1]. The bacterium occurs naturally in soil and commonly affects domestic and wild animals around the globe. Livestock workers and household animal owners are directly exposed to anthrax disease due to their close interactions with animals [2]. Generally, rearing animals either for consumption or commercial purposes is a major source of livelihood in many low- and middle-income countries (LMICs) with increased exposure and susceptibility to the infectious agent and disease [3, 4].

Anthrax is endemic in many parts of Africa, with recurrent outbreaks reported in several regions of the continent [3, 5]. The major risk factors for outbreaks and spread of anthrax between species include environmental contamination and exposure through grazing or ingestion [5, 6]. Outbreaks tend to occur mostly during the dry season, affecting humans, livestock, and wildlife [3, 5]. For instance, about 67% of wildlife anthrax outbreaks in Kenya occurred during the dry season [3]. Similarly, most of the human anthrax cases recorded in Tanzania’s hotspot regions were diagnosed during the dry season [7].

Nigeria is the most populous country in Africa, with an estimated population of 206 million in 2022 [8]. Agriculture is the country’s mainstay, having one of the highest populations of livestock (19.5 million cattle, 72.5 million goats, 41.3 million goats) in the continent [9]. Livestock rearing, including pastoralism, is very prominent, and most households have close contact with domestic animals. Nonetheless, awareness of the public health implications of zoonotic diseases is poor [10, 11]. Anthrax is ranked as one of the first five priority zoonoses in Nigeria [12].

Similar to other LMICs, the burden of zoonotic diseases, including anthrax, in Nigeria is often underestimated partly due to poor awareness, inadequate preventive measures, and non-existence of active surveillance mechanism, which increases the risk of their spread [13, 14]. The knowledge of causative agents, modes of transmission, clinical manifestations, and potential consequences of zoonotic diseases are key for effective control. As a zoonotic and vaccine-preventable disease, adequate vaccination of susceptible animal populations reduces the risk of transmission to humans. Hence, reducing exposure to infected animals or their by-products and the control of animal anthrax reduce human risk. While the primary control measure for anthrax in animal is annual preventive vaccination, control measures, such as ring vaccination, proper carcass disposal, isolation and quarantine of new or affected animals, could reduce the spread of the disease, especially during outbreaks [15]. In addition, re-vaccination of animals on antibiotic regimens is very pertinent to ensure proper protection [15, 16].

The recent outbreak of anthrax in Nigeria is a matter of public health concern, especially considering the weak surveillance in the country. Nigeria activated its emergency preparedness and response activities to prevent the incursion of the disease into the country in response to the confirmation of the outbreak in Ghana on the 1^st^ of June, 2023. The Federal Ministry of Agriculture, through the Department of Veterinary and Pest Control Services, activated the National Anthrax Technical Working Group (NTWG). The NTWG is a multisectoral/multidisciplinary committee comprising stakeholders from the human, animal and environmental health sectors and partners. The activities included creating an increased awareness of the outbreak among the Directors of Veterinary Services and the State and Federal Epidemiology officers in all 36 States and the Federal Capital Territory. Media houses also intensified their efforts to increase awareness among the general public about the outbreak of anthrax and the necessary precautions to be taken. Also, the necessary steps and reporting channels in the event of a suspect were defined.

Despite the prompt response and preventive measures, on the 16^th^ of July 2023, the Niger State Ministry of Agriculture, with support from the Federal Ministry of Agriculture and Food Security (FMAFS), Nigeria Center for Disease Control (NCDC) and partners, confirmed an outbreak of anthrax in a farm in Suleja, Niger State, North Central Nigeria. In addition, Lagos State reported a suspected outbreak. Since the declaration of the outbreak, several activities were initiated, including intensive risk communication activities and heightened public awareness campaigns on anthrax at abattoirs, livestock markets and hunting communities. Also, surveillance activities were intensified at national and international control posts, abattoirs, cattle markets and other livestock and “bush meat” markets. Considering this background, this study sought to assess the knowledge, perception, and preventive practices of livestock workers and household animal owners toward anthrax in Nigeria and the associated socioeconomic impacts of the disease.

## Materials and methods

### Study Area

Nigeria is situated in the western part of Africa. It is bordered in the north by the Niger and Chad Republics, in the east by Cameroon, in the west by the Benin Republic, and in the south by the Gulf of Guinea and the Atlantic Ocean. Nigeria has a land mass that spans an area of 923,769 square kilometres and has a population of over 230 million people [8]. On the globe, Nigeria lies between 4 °15’- 13 15’North latitude and longitude 2 °-40’-14 °-45 East [18]. The climate in Nigeria varies largely, with tropical rainforests and an average annual rainfall of 1500-2000 millimetres found in the southernmost region of the country. The entire land mass between the far north and far south of the country is savannah, with an average annual rainfall between 500 – 1500 millimetres [17].

Agriculture, including livestock farming, is the major occupation, with most households having regular contact with animals. The country has several food animal slaughter facilities across the states and Local Government Areas. Aside from serving as meat processing facilities, these areas constitute critical points for the exposure of humans to zoonotic diseases. Seven out of the 36 states in Nigeria were enrolled for the study: these included, Niger, Lagos, Ogun, Oyo, Benue, Ebonyi and Bayelsa (Figure 1). The choice of the states was informed by having reported anthrax outbreak, being a contiguous state to reporting states and presence of livestock activities and ease of logistics.

**Fig 1:**
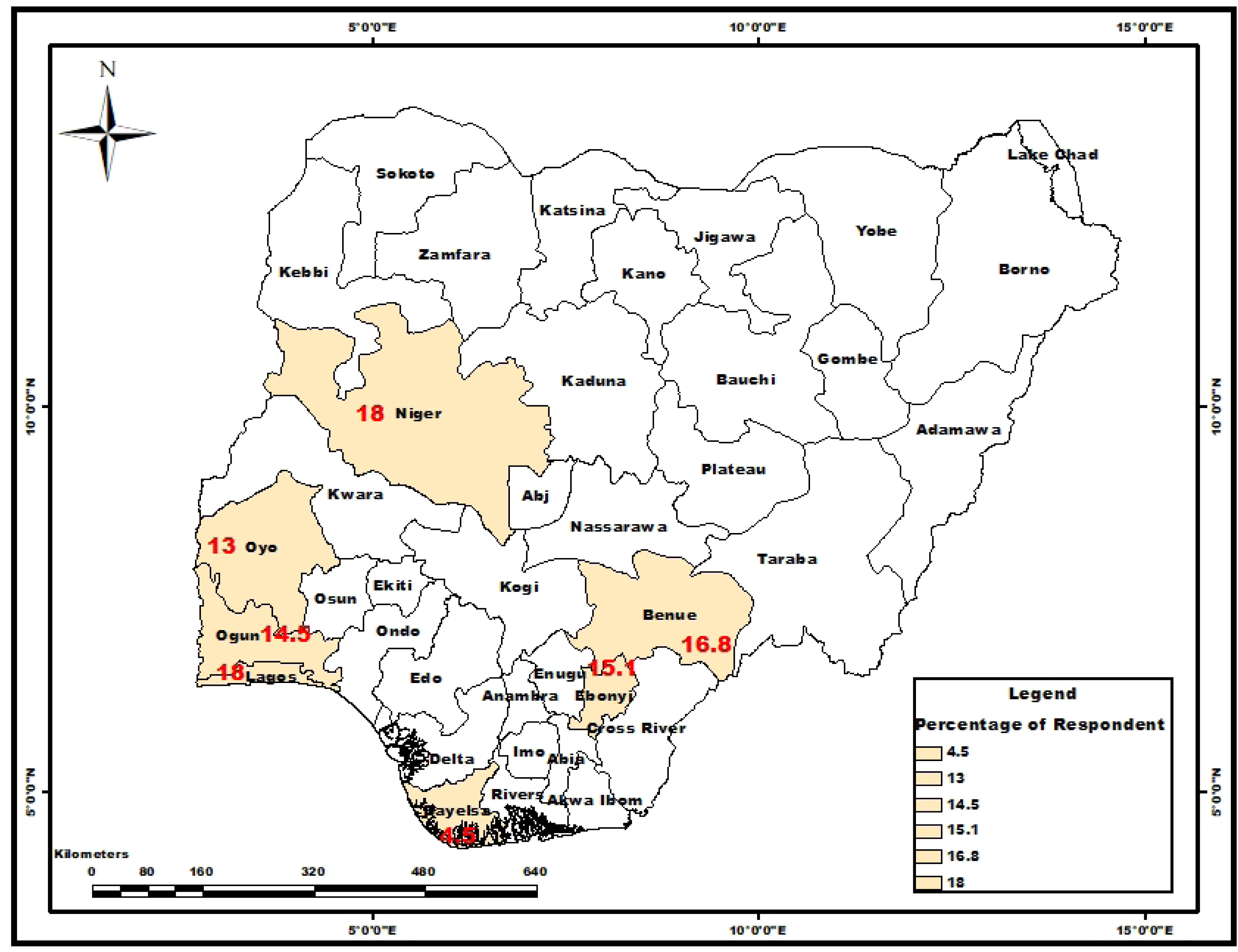
Map of Nigeria showing study sites, with percentage of respondents interviewed in each state. Source: Department of Geography, University of Ibadan; https://Grid3.org.

### Study Design

Using a cross-sectional design, relevant data were obtained to assess the knowledge, perception, and preventive practices of livestock workers and household animal owners toward anthrax, as well as the risk of exposure and socioeconomic impacts associated with the disease. The study was carried out from August to September 2023. Data were obtained using REDCap (Research Electronic and Data Capture) software.

### Study Population

The study population included livestock workers (meat sellers, livestock traders, pastoralists) and household animal owners.

### Sample Size Determination and sampling

A total of 1025 participants were interviewed for this study. This number was determined using the formula for survey [19] at a 95% confidence level, a 5% level of precision, and assuming a 50% expected proportion of community members with knowledge of the cause/symptom or mode of transmission of anthrax. The estimated sample was adjusted for design effect of 2.0 and a 25% non-response rate. Cluster sampling was adopted and all consenting livestock workers and households with animals were involved in the study.

### Eligibility Criteria

#### Inclusion Criteria

Males and females aged 18 years and above, who have previously or presently owned, reared, or sold livestock animals were involved in the study.

#### Exclusion Criteria

Livestock workers and household animal owners who refused to give their consent or who were unable to give a response due to incapacitation or communication problems were excluded from the study.

### Data Collection

A semi-structured, interviewer-assisted questionnaire was used for data collection. The questionnaire consisted of ten sections, including respondents’ information, sociodemographic characteristics, animal ownership, and the risk of exposure to anthrax and other zoonoses. Other variables included awareness about anthrax, knowledge of on anthrax, perception about anthrax, practices towards anthrax prevention, history of anthrax vaccination, and socioeconomic impacts of anthrax. Ten and six-item questions were used to assess the knowledge and perception on anthrax, respectively.

The demographic section of the questionnaire was developed from the Nigeria Demographics and Health Survey (NDHS). Other sections were developed from similar studies [4, 20] and reviewed by experts in the field. Fifteen research assistants who understand the local dialects of the study sites were recruited for the study. The research assistants were trained in the use of the REDCap software and the identification of common external symptoms of anthrax disease in animals and humans. The questionnaire was translated into the common local dialects and back-translated into the English language to ensure the original meanings were retained.

### Reliability and Validity

A pretest was conducted among 10 participants with similar characteristics in a different location from the study area. The questionnaire was adjusted accordingly after the pretest. The consistency indicator (Cronbach’s alpha coefficient) for the questions used to assess the knowledge and perception on anthrax were 0.90 and 0.76, respectively.

### Data Analysis

Data were analysed using the Statistical Package for the Social Sciences (SPSS) version 29. The demographic characteristics of the respondents were analysed using descriptive statistics and were presented using frequencies and percentages. Each item on the risk of exposure to anthrax was graded as 0, 2 and 4 representing low, moderate and high risk, respectively. The maximum obtainable score was 34, with less than 8 representing low risk, while 8 to 16 was categorized as moderate risk and greater than 16 as high risk. The questions on knowledge were allocated unequal points based on the weight of each question. The maximum point obtainable was 72. Respondents with score of 36 and above were categorized as having good knowledge. The perception of the respondents towards anthrax was calculated using a 4-point Likert scale with 1 point allocated to strongly disagree, and 4 points to strongly agree. The maximum obtainable score was 24. Respondents with aggregate score of ≥ 15 were categorized as having positive perception.

The preventive practices against anthrax and the socioeconomic impacts of anthrax on the respondents were analyzed using frequencies and percentages. The associations between the independent variables (sociodemographic factors) and risk of exposure, as well as knowledge were determined accordingly using Chi-square test. All tests were two-tailed and statistical significance was set 5% level.

### Ethical Consideration

Ethical approval was obtained from the University of Ibadan/University College Hospital Institutional (UI/UCH) Ethics Review Committee (UI/UCH/22/0305). Informed consent was obtained from respondents, after which the questionnaires were administered. Due to the documented low literacy level among the study population, endorsement of the consent form included signing by those who could and thumbprint or verbally. The respondents were assured of the voluntary nature of the study and the right to decline or withdraw. This action will not be held against them, and they were assured that there will be no adverse consequences. The data collected were strictly confidential and stored on a password-protected computer. Identifiers such as names and addresses were excluded.

## Results

### Sociodemographic characteristics of participants

A total of 1025 respondents from seven states in Nigeria participated in the study. Table 1 shows the sociodemographic characteristics of the respondents. The mean age of the respondents was 46.7±11.67 years. The respondents were from Lagos (18.0%), Niger (18.0%), Benue (16.8%), Ebonyi (15.1%), Ogun (14.5%), Oyo (13.0%) and Bayelsa (4.5%) states. The majority of the respondents were males (75.6%) and married (79.4%). About half were Moslems (51.3%) while only (36.6%) had up to secondary level education, and 3.0% of the respondents had no formal education. The majority (86.2%) of the respondents were employed and were livestock workers (69.3%).

**Table 1.**
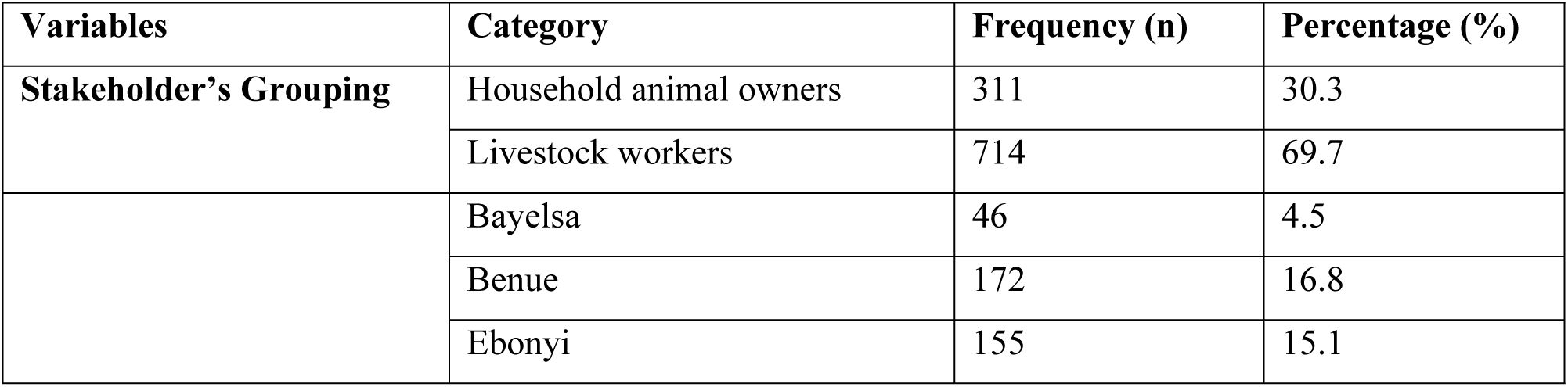
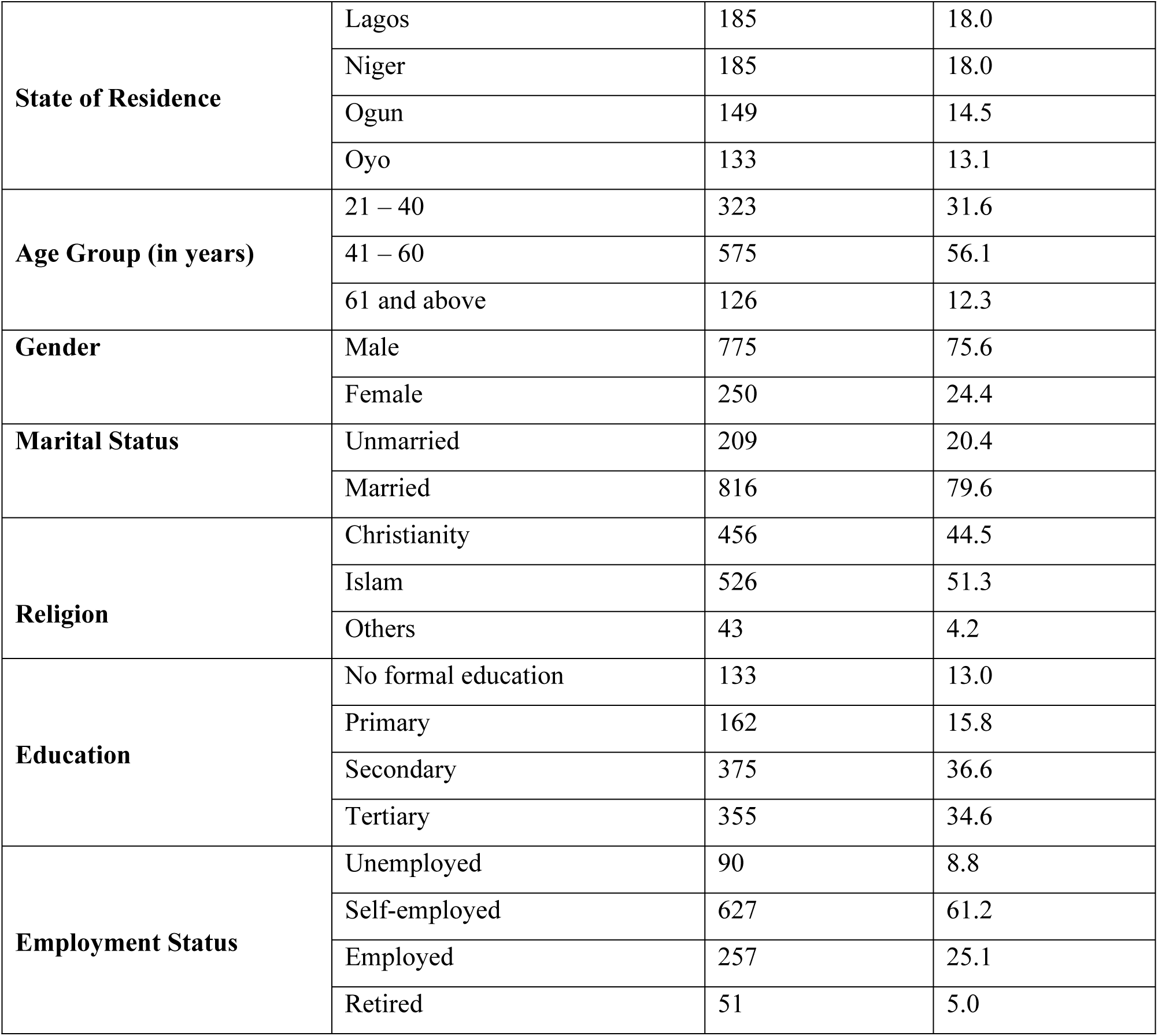
Sociodemographic characteristics of the respondents.

### Respondents’ risk of exposure to anthrax

Most 773 (75.4 %) of the respondents had low risk of exposure to anthrax (Table 2). Exactly 67.6% of the respondents indicated having contact with the soil through day-to-day activities. Only about a third (39.0%) of the respondents said they would report an animal death of unknown cause to a veterinary centre; 38.8% indicated discarding the animal by burying it, with only 6.6% and 2.0% either eating or selling it, respectively. The majority (86.6%) reported that they would go for vaccination to protect themselves from anthrax, while most (91.6%) of the participants agreed that it was important to take their animals for vaccination. Up to 68.7% of the respondents indicated they would be concerned if the body of a dead animal did not become stiff. More than half (61.6%) of the respondents said they would call a veterinary doctor if they saw frank, unclotted blood coming out of the natural openings of their animals, while almost a quarter (23.3%) said they would throw the animal away (Table 2).

**Table 2.** Respondents’ risk of anthrax disease (n = 1025)

| **Variable** | **Frequency**<br>**(n)** | **Percentage**<br>**(%)** |
| --- | --- | --- |
| **Do your day-to-day activities involve contact with the soil?** |  |  |
| Yes | 693 | 67.6 |
| No | 332 | 32.4 |
| **What will you do if you see an animal that dies of an unknown cause?** |  |  |
| Sell it | 21 | 2.0 |
| Eat at home | 68 | 6.6 |
| Throw it away | 282 | 27.5 |
| Discard by burying | 398 | 38.8 |
| Report to veterinary | 400 | 39.0 |
| Others | 52 | 5.1 |
| **Will it be of concern to you if the dead animals' bodies did not become stiff?** |  |  |
| Yes | 704 | 68.7 |
| No | 321 | 31.3 |
| **Will it be of concern to you if you see an animal with unclotted blood coming out of its natural opening?** |  |  |
| Yes | 905 | 88.3 |
| No | 120 | 11.7 |
| **What will you do if you see an animal with unclotted blood coming out of its natural opening?** |  |  |
| Quickly slaughter for consumption | 52 | 5.1 |
| Slaughter for sale? | 98 | 9.6 |
| Clean the blood? | 63 | 6.1 |
| Call a veterinarian? | 629 | 61.4 |
| Throw it away? | 238 | 23.2 |
| Others? | 51 | 5.0 |
| **Is vaccination of animals important in preventing animal diseases?** |  |  |
| Yes | 939 | 91.6 |
| No | 86 | 8.4 |
| **Would you go for vaccination as a means of preventing yourself from diseases?** |  |  |
| Yes | 888 | 86.6 |
| No | 137 | 13.4 |
| **Risk of exposure to anthrax** |  |  |
| Low | 773 | 75.4 |
| Moderate | 244 | 23.8 |
| High | 8 | 0.8 |

### The awareness of respondents about anthrax disease

About 71.2% reported they have heard of anthrax previously. The main sources of respondents’ information about anthrax were radio (38.7%), community health workers/veterinarians (30.2%), and television (25.9%). The social media (22.6%), friends/neighbour/family (18.5%), health professionals (6.7%), print media (6.3%), posters (5.7%), religious leaders (2.9%) and billboards (1.8%) were the other sources of information about anthrax. The majority (67.3%) of the respondents were aware of the recent anthrax outbreak in Nigeria, but only 2.4% and 6.8% had seen a person or an animal with anthrax disease, respectively.

### Knowledge of respondents about anthrax disease

The majority (58.6%) of the respondents had good knowledge of anthrax disease (Mean + SD: 38.75+14.18). The respondents agreed that consumption of contaminated meat (43.6%) and direct contact with infected animals (44.6%) are sources of disease transmission to humans. The common symptoms of anthrax disease in humans identified by the respondents were skin rash or sores/wounds (38.3%), fever (35.5%), fatigue (23.4%), respiratory distress (28.4%) and muscle aches and pain (21.3%). Also, bleeding from natural openings (51.4%), sudden death (41.0%), and unclotted, dark red blood (38.6%) were the symptoms of anthrax in animals indicated by the respondents. In terms of respondents’ knowledge of anthrax prevention, vaccination of livestock (49.7%), hand washing after handling animals (37.0%), and isolating and treating infected animals promptly (36.9%) were the preventive measures highlighted (Table 3).

**Table 3.**
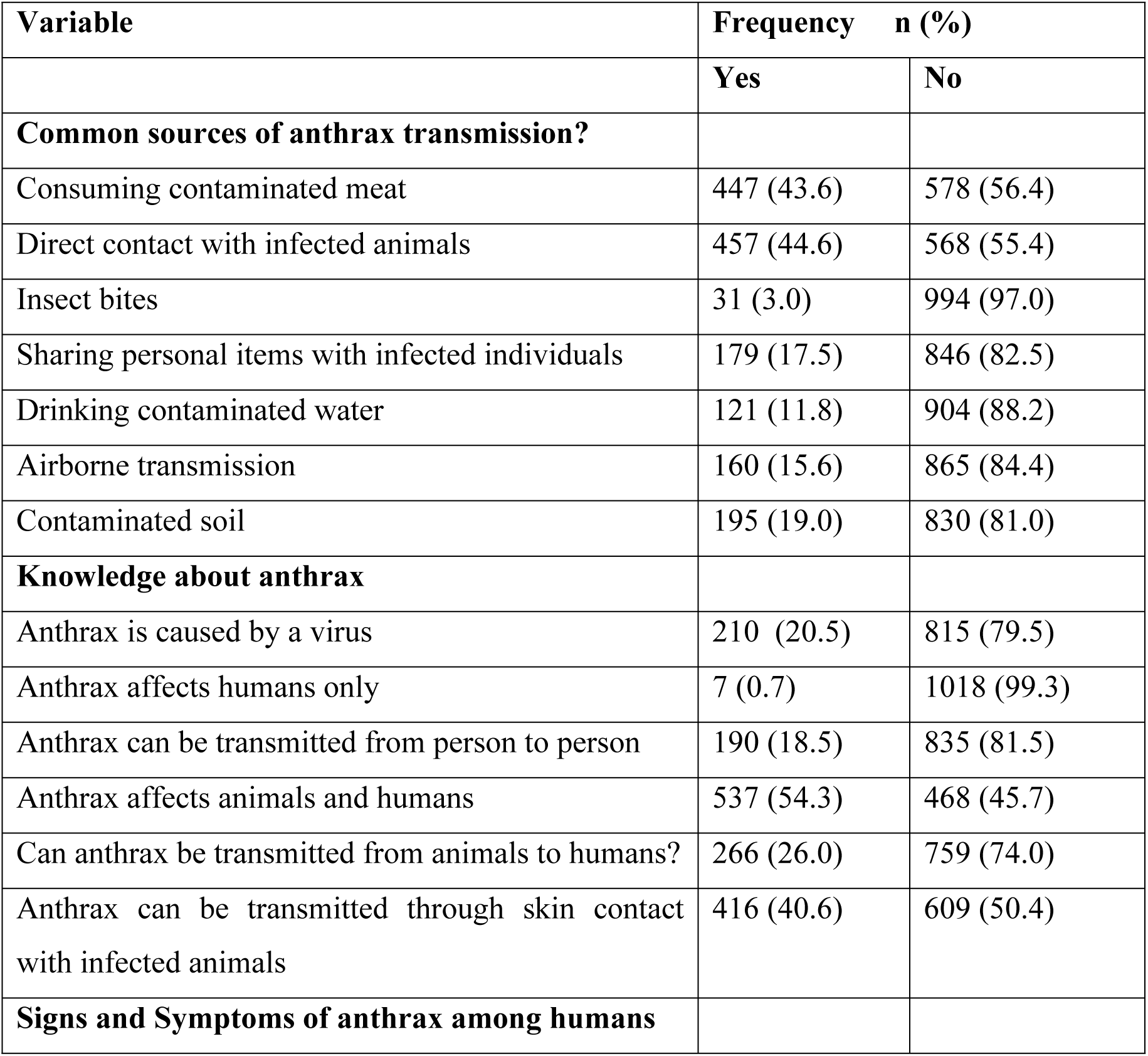
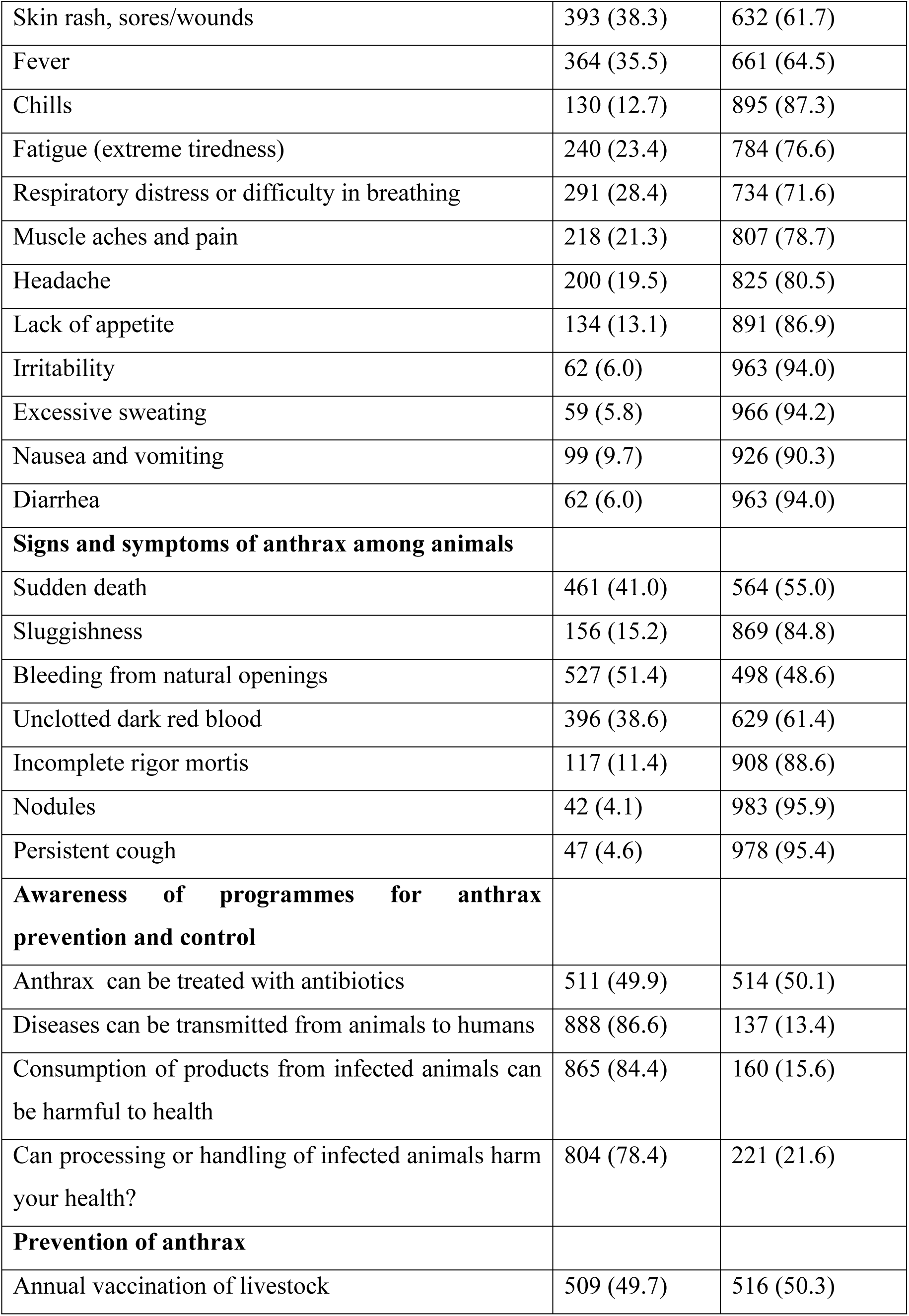
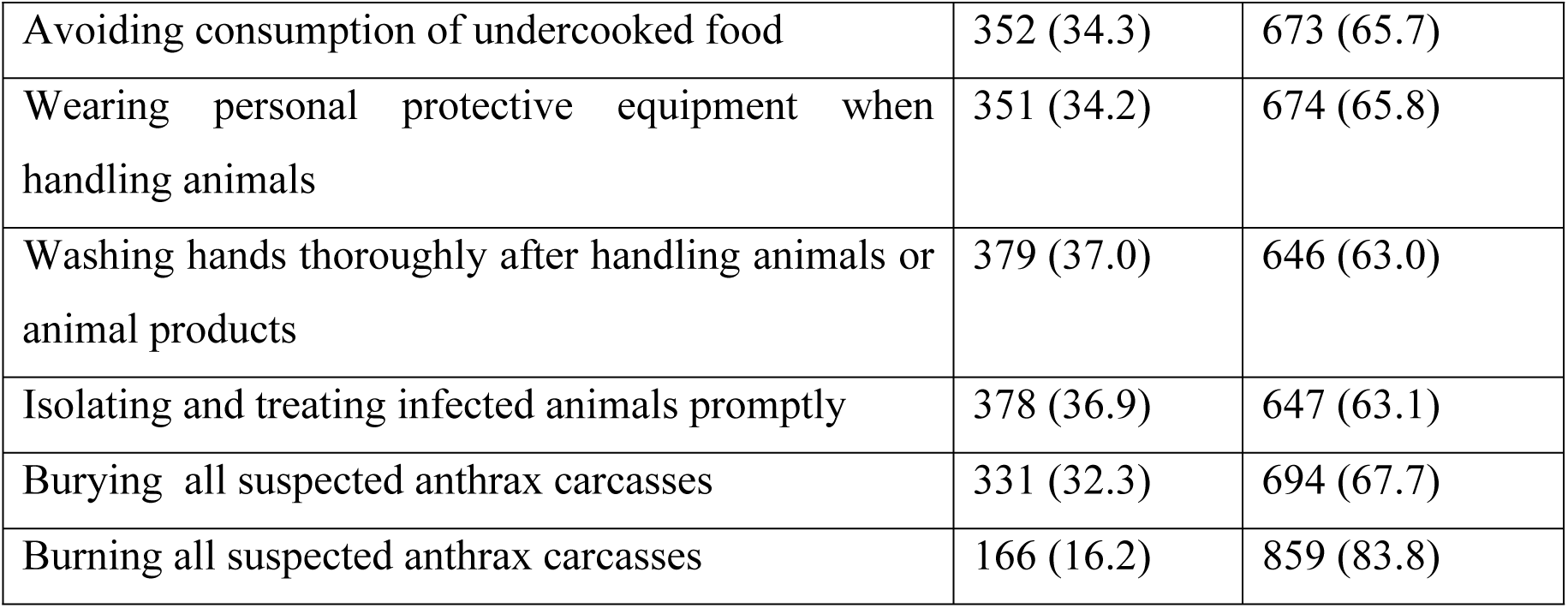
Respondent’s knowledge of anthrax disease (n = 1025)

### Perception of respondents about anthrax disease

The majority (79.9%) of the respondents showed a positive perception towards anthrax disease, the mean±SD score being 11.62+3.42. Up to 67.6% believed that anthrax was a significant threat to human and animal health. More than half (54.8%) of the respondents were very concerned about the possibility of an anthrax outbreak in their area, and 74.0% considered educating the public about anthrax as very important. However, more than half (58.0%) of the respondents did not think that anthrax was a serious disease in animals. In comparison, only 9.0% considered it a disease of serious public health implication in humans (Table 4).

**Table 4.** Respondents’ perception towards anthrax disease (n = 1025)

| **Variable** | **Frequency (n)** | **Percentage (%)** |
| --- | --- | --- |
| **Do you believe that anthrax is a significant threat to human and animal health in Nigeria?** |  |  |
| Yes | 693 | 67.6 |
| No | 332 | 32.4 |
| **In your opinion, how serious a disease is anthrax in animals in your area?** |  |  |
| Very serious | 149 | 14.5 |
| Somewhat serious | 281 | 27.4 |
| Not very serious | 595 | 58.1 |
| **In your opinion, how serious a disease is anthrax in humans in your area?** |  |  |
| Very serious | 92 | 9.0 |
| Somewhat serious | 151 | 14.7 |
| Not very serious | 782 | 76.3 |
| **How would you rate your knowledge of anthrax** |  |  |
| Very knowledgeable | 217 | 21.2 |
| Moderately knowledgeable | 268 | 26.2 |
| Slightly knowledgeable | 351 | 34.2 |
| Not at all knowledgeable | 189 | 18.4 |
| **How concerned are you about the possibility of an anthrax outbreak in your area?** |  |  |
| Very concerned | 583 | 56.9 |
| Moderately concerned | 150 | 14.6 |
| Slightly concerned | 168 | 16.4 |
| Not at all concerned | 124 | 12.1 |
| **In your opinion, how important is it to educate the public about anthrax?** |  |  |
| Very important | 768 | 74.9 |
| Moderately important | 110 | 10.7 |
| Slightly important | 83 | 8.1 |
| Not at all important | 64 | 6.2 |

### Respondents’ preventive practices towards anthrax

About half (50.2%) of the respondents practised a mix of free range and zero grazing, and about a third (35.2%) bought commercial fodder to feed their animals. However, only 22.0% had ever taken any measure to protect themselves or their animals from anthrax infection. In all, 0.7% of the respondents had a history of anthrax infection in their animals, while 0.8% of them had had a family member infected with anthrax (Table 5).

**Table 5.** Respondents’ preventive practices towards anthrax (n = 1025)

| Variable | Frequency<br>(n) | Percentage<br>(%) |
| --- | --- | --- |
| **What animal husbandry do you practice?** |  |  |
| Zero grazing | 193 | 18.8 |
| Free range | 222 | 21.7 |
| Mixed free range and zero grazing | 515 | 50.2 |
| Others | 95 | 9.3 |
| **Where do you get fodder for your animals?** |  |  |
| I graze in the field | 335 | 32.7 |
| I cut and carry fodder | 230 | 22.4 |
| I buy commercial fodder | 361 | 35.2 |
| Others | 99 | 9.7 |
| **Have you ever taken any preventive measures to protect yourself or your livestock from anthrax?** |  |  |
| Yes | 226 | 22.0 |
| No | 799 | 78.0 |
| **Has any of your animals died suddenly before** |  |  |
| Yes | 131 | 12.8 |
| No | 894 | 87.2 |
| **Has anthrax infected your animal** |  |  |
| Yes | 7 | 0.7 |
| No | 1018 | 99.3 |
| **Has any member of your family suffered from anthrax** |  |  |
| Yes | 8 | 0.8 |
| No | 1017 | 99.2 |
| **How did family members contract it?** |  |  |
| Eating dead animal | 4 | 50 |
| Carrying hide from a dead animal | 0 | 0 |
| Carrying meat from the dead animal | 1 | 12.5 |
| Others | 3 | 37.5 |
| **What actions did you take?** |  |  |
| Took the person to the nearest health facility | 2 | 25.0 |
| Bought medicine from a chemist | 0 | 0 |
| Took the person to a traditional healer | 3 | 37.5 |
| Others | 37.5 | 37.5 |

### History of Anthrax Vaccination

Only a few (27.7%) of the respondents knew about the anthrax vaccination programme for livestock in their area. Also, only 19.7% had encountered or heard about anthrax outbreaks or cases in their community or nearby areas, out of which 82.2% heard about the cases less than six months ago. The majority (84.7%) of the respondents who had encountered or heard about anthrax cases in their community confirmed veterinary intervention regarding the cases, and 80.2% of them said animal vaccination was embarked upon during the period. Regarding the frequency of animal vaccination, 32.9% reported that the veterinary personnel were always available to vaccinate in their area, while 12.5% indicated that the vaccination was done yearly in their area. Slightly above half (55.1%) of the respondents said they often take their animals for vaccination. In comparison, 32.6% of the respondents who do not often take their animals for vaccination said it was because they were not informed of any vaccination (Table 6).

**Table 6.** History of anthrax vaccination.

| **Variable** | **Frequency (n)** | **Percentage (%)** |
| --- | --- | --- |
| **Do you have any knowledge of the anthrax vaccination program for livestock in your area?** |  |  |
| Yes | 275 | 27.7 |
| No | 718 | 72.3 |
| **Have you encountered or heard of any anthrax cases in your community or nearby areas?** |  |  |
| Yes | 202 | 19.7 |
| No | 823 | 80.3 |
| **If yes, when did you encounter or hear about it** |  |  |
| Less than 6 months ago | 166 | 82.2 |
| One year ago | 12 | 5.9 |
| More than one year ago | 4 | 2.0 |
| Others | 20 | 9.9 |
| **Was there any response from the veterinary department in your area** |  |  |
| Yes | 171 | 84.7 |
| No | 31 | 15.4 |
| **Was there vaccination during the period** |  |  |
| Yes | 162 | 80.2 |
| No | 40 | 19.8 |
| **If yes, were all animals vaccinated** |  |  |
| Yes | 146 | 72.3 |
| No | 56 | 27.7 |
| **If anthrax vaccination programs for livestock are available in your area, would you be willing to participate** |  |  |
| Yes | 832 | 81.2 |
| No | 193 | 18.8 |
| **How often is animal vaccination done in your area?** |  |  |
| Twice a year | 25 | 2.4 |
| Once a year | 128 | 12.5 |
| The veterinary personnel are always available to vaccinate | 337 | 32.9 |
| Never available | 336 | 32.7 |
| Other | 200 | 19.5 |
| **Do you take your animals for vaccination?** |  |  |
| No | 460 | 44.9 |
| Yes | 565 | 55.1 |
| **What prompts you to take your animal for vaccination** |  |  |
| To protect animals | 312 | 55.2 |
| To protect humans | 19 | 3.4 |
| Because others do so | 3 | 0.5 |
| Because it is a requirement | 227 | 40.2 |
| Others | 4 | 0.7 |
| **If you do not always take your animals for vaccination, what are the reasons?** |  |  |
| No vet services | 13 | 2.8 |
| Financial difficulties | 30 | 6.5 |
| Don't get informed when it occurs | 150 | 32.6 |
| The vaccination centre is far | 20 | 4.3 |
| Others | 247 | 53.7 |
| **In your opinion, would vaccination of animals help to prevent anthrax** |  |  |
| Yes | 863 | 84.2 |
| No | 162 | 15.8 |

### The Socioeconomic impact of anthrax

Most (99.1%) of the respondents said that anthrax disease had never affected their livestock, farm, or household animals, while 79.7% said that the current anthrax outbreak had a minor effect on their business. Again, only 23.8% of the respondents agreed that restrictions or regulations imposed during the anthrax outbreak affected their livestock-related activities (Table 7).

**Table 7.** Socioeconomic impact of anthrax.

| Variable | Frequency (n) | Percentage (%) |
| --- | --- | --- |
| **Would you be willing to report suspected anthrax cases to the relevant authorities if you come across them?** |  |  |
| Yes | 952 | 92.9 |
| No | 73 | 7.1 |
| **Has anthrax ever affected your livestock, farm, or household animals** |  |  |
| Yes | 9 | 0.9 |
| No | 1016 | 99.1 |
| **To what extent has the current anthrax outbreak affected your business** |  |  |
| High | 10 | 1.0 |
| Moderate | 198 | 19.3 |
| Low | 817 | 79.7 |
| **Do any restrictions or regulations imposed during an anthrax outbreak affect your livestock-related activities** |  |  |
| Yes | 244 | 23.8 |
| No | 781 | 76.2 |
| **Are you afraid or worried that you might go out of business due to the current anthrax outbreak?** |  |  |
| Yes | 416 | 40.6 |
| No | 609 | 59.4 |
| **Have you ever sought financial or medical assistance from the government or other organisations due to anthrax-related issues?** |  |  |
| Yes | 8 | 0.8 |
| No | 1017 | 99.2 |

### Factors associated with knowledge of anthrax among livestock workers and household animal owners in Nigeria

There was a significant association between the respondents’ knowledge of anthrax and their level of education (p=0.000), awareness level (p=0.000), perception (p=0.000) and risk level (p=0.000) (Table 8). The respondents with education below secondary level were less likely (aOR: 0.69; 95%CI: 0.51–0.94) to have good knowledge of anthrax than those with secondary education and above. The respondents who were aware of anthrax were about 5 times more likely (aOR: 5.35; 95%CI: 3.87–7.39) to have good knowledge of anthrax than those without awareness of the disease. Respondents with positive perception about anthrax were about twice more likely (aOR: 2.00; 95%CI: 1.38 – 2.90) to have good knowledge of anthrax than those with negative perception (Table 8).

**Table 8.** Factors associated with knowledge of anthrax among livestock workers and household animal owners in Nigeria.

|  | **Knowledge of Anthrax** |  | **OR (95% CI)** | **AOR (95% CI)** | **P-Value** |
| --- | --- | --- | --- | --- | --- |
| **Stakeholders Grouping** | **Good (n = 601)** | **Poor (n = 424)** |  |  |  |
| Household animal workers | 188 | 123 | 0.898 (0.684 – 1.178) |  | 0.436 |
| Livestock workers | 413 | 301 | ref |  | - |
| **Sex** |  |  |  |  |  |
| Male | 450 | 325 | 0.908 (0.679 – 1.214) |  | 0.514 |
| Female | 151 | 99 | ref |  |  |
| **Age Group** |  |  |  |  |  |
| 46 and above | 325 | 235 | 1.056 (0.823 – 1.356) |  | 0.669 |
| Below 46 | 276 | 189 |  |  |  |
| **Marital Status** |  |  |  |  |  |
| Married | 477 | 339 | 1.037 (0.761 – 1.412) |  | 0.819 |
| Unmarried | 124 | 85 | ref |  |  |
| **Level of Education** |  |  |  |  |  |
| Less than secondary school | 133 | 162 | 0.46 (0.35 – 0.60) | 0.69 (0.51 – 0.94) | 0.02 |
| Secondary school and above | 468 | 262 | ref | ref |  |
| **Employment Status** |  |  |  |  |  |
| Unemployed | 87 | 54 | 0.862 (0.599 – 1.242) |  | 0.426 |
| Employed | 514 | 370 | ref |  |  |
| **Awareness** |  |  |  |  |  |
| Yes | 523 | 207 | 7.03 (5.18 – 9.53) | 5.35 (3.87 – 7.39) | <0.001 |
| No | 78 | 217 | ref | ref |  |
| **Perception Level** |  |  |  |  |  |
| Positive | 535 | 284 | 4.00 (2.88 – 5.54) | 2.00 (1.38 – 2.90) | <0.001 |
| Negative | 66 | 140 | ref | ref |  |
| **Exposure Risk Level** |  |  |  |  |  |
| Low | 484 | 289 | 1.93 (1.45 – 2.58) |  | <0.001 |
| Moderate & High | 117 | 135 | ref |  |  |

Further, Table 9 shows the relationship between the respondents’ knowledge and their educational level, awareness, perception and risk of exposure. Respondents who were married had lower odds of exposure risk (aOR: 0.57 95%CI: 0.28 – 0.83) to anthrax than those who were unmarried. Respondents with education below secondary level had higher odds of exposure risk (aOR: 1.85; 95%CI: 1.33 – 2.56) than those with education above secondary level. Respondents who were aware of anthrax had lower odds of exposure risk (aOR: 0.32; 95%CI: 0.23 – 0.45) than those without awareness. Respondents with positive perception about anthrax had lower odds of exposure risk (aOR: 0.47; 95%CI: 0.33 – 0.68) than those with negative perception.

**Table 9.** Factors associated with risk of exposure to anthrax among livestock workers and household animal owners in Nigeria.

|  | Exposure risk to Anthrax |  | OR (95% CI) | AOR (95% CI) | P-Value |
| --- | --- | --- | --- | --- | --- |
| Stakeholders Grouping | Moderate & High (n = 252) | Low (n = 773) |  |  |  |
| Household animal workers | 78 | 233 | 0.96 (0.71 – 1.31) |  | 0.81 |
| Livestock workers | 174 | 540 | ref | - |  |
| **Sex** |  |  |  |  |  |
| Male | 180 | 595 | 0.748 (0.543 – 1.030) |  | 0.075 |
| Female | 72 | 178 | ref |  |  |
| **Age Group** |  |  |  |  |  |
| 46 and above | 131 | 429 | 0.868 (0.653 – 1.154) |  | 0.331 |
| Below 46 | 121 | 344 | ref |  |  |
| **Marital Status** |  |  |  |  |  |
| Married | 181 | 635 | 0.65 (0.52 – 0.83) | 0.57 (0.28 – 0.83) | <0.01 |
| Unmarried | 71 | 138 | Ref |  |  |
| **Level of Education** |  |  |  |  |  |
| Less than secondary school | 112 | 183 | 2.58 (1.91 – 3.48) | 1.85 (1.33 – 2.56) | <0.001 |
| Secondary school and above | 140 | 590 | Ref |  |  |
| **Employment Status** |  |  |  |  |  |
| Unemployed | 31 | 110 | 0.85 (0.55 – 1.30) |  | 0.60 |
| Employed | 221 | 663 | Ref |  |  |
| **Awareness** |  |  |  |  |  |
| Yes | 114 | 616 | 0.21 (0.16 – 0.29) | 0.32 (0.23 – 0.45) | <0.001 |
| No | 138 | 157 | ref |  |  |
| **Perception Level** |  |  |  |  |  |
| Positive | 156 | 663 | 0.33 (0.27 – 0.50) | 0.47 (0.33 – 0.68) | <0.001 |
| Negative | 96 | 110 | ref |  |  |
| **Knowledge Level** |  |  |  |  |  |
| Good | 117 | 484 | 0.52 (0.39 – 0.69) |  | <0.001 |
| Poor | 135 | 289 | ref | - | - |

## Discussion

This study investigated the knowledge and perception of anthrax as well as the socioeconomic impacts of the disease among populations at risk (livestock owners and household animal owners) in Nigeria. The sociodemographic characteristics of the respondents in this study provide important insights into the wide age range profile of individuals engaged in livestock-related activities and their potential exposure to anthrax. Such diversity in age groups underscores the need for tailored educational campaigns that are accessible and relevant to both young and old. Again, the predominance of male participants in this study aligns with the traditional gender roles often associated with livestock farming and animal husbandry [21]. The involvement of physical tasks and outdoor work may explain the higher male participation. Similar to the findings by Sitali *et al*. [22], this study revealed a significant association between the educational level of the respondents and their knowledge of anthrax. Similar findings were reported from Zambia [22] and Ethiopia [20] both indicated good awareness about anthrax among respondents. This high level of awareness may be connected to the sensitisation campaigns by the Nigerian government consequent upon the recent outbreak of anthrax [23]. The most common sources of information were radio, community health workers/veterinarians and television, which highlight the role of mass media and healthcare professionals in disseminating information about anthrax and other health-related issues.

Regarding the disposition of the respondents to vaccination, the majority reported that they would go for vaccination to protect themselves from anthrax and that it was important to take their animals for vaccination. This finding is encouraging since routine vaccination policy is a major mitigation and prevention strategy against anthrax [7].

Further, our findings reveal that just over half of the respondents had good knowledge of anthrax. This is lower compared to the findings of Dutta et al. [4], where 62.73% of the livestock farmers in selected rural areas of Bangladesh had good knowledge about anthrax. It is also lower when compared with 64% earlier reported [24] among livestock farmers and consumers in Southern Ethiopia. However, our finding is similar to the 53.8% reported among selected households in villages of Southern Kenya [25]. This level is low considering the recent sensitisation campaigns by the Nigeria government against anthrax and the fact that the respondents, assessed in this study, were an at-risk population who were expected to be abreast of health-related issues associated with animals. This therefore, suggests that there are key knowledge gaps among the respondents that interventional programmes should be focused on. Such gaps should include annual vaccination of animals, training on recognition of common signs of anthrax in animals, and knowledge of transmission routes of anthrax. Hence, subsequent sensitization and educational programmes should take into consideration such knowledge gaps from the planning phase of the intervention. Notably, in this study, the knowledge level of the respondents was significantly associated with their education, awareness, perception and risk of anthrax. Consequently, programmes geared towards increasing knowledge levels on anthrax should include activities that could promote education and awareness as well as positively influence the perception of the livestock workers and household animal owners in Nigeria.

Moreover, an assessment of the perception of the respondents shows that the majority had a positive perception towards anthrax disease and believed that anthrax was a significant threat to human health. This finding is suggestive of the readiness of respondents to embrace educational programmes about anthrax. As such, achieving behavioural change in a positive direction could be enhanced. It is however worrisome that more than half of the respondents did not think that anthrax was a serious disease in animals. This is of great concern, since anthrax is a zoonotic disease that is mostly transmitted from animals to humans. Livestock owners are supposed to be the first gatekeepers.

Again, the respondents engaged in certain practices that could expose their animals to anthrax infection as well as jeopardise human health, such as open grazing, and cutting of fodder for animal feeding. However, a majority reported not taking any preventive measures to either protect themselves or their animals against anthrax. As reported, ingestion is the most natural route of infection, resulting in fatal gastrointestinal anthrax with massive replication of bacilli in all organ tissues [26]. Considering the poor handling of dead carcasses, as observed in this study, unguarded open grazing or harvesting of fodder might constitute a health threat to the animals. This is because decomposition of animal carcasses and exposure to air (oxygen) trigger a differentiation process that releases spores [26]. While they do not replicate, spores persist in the environment until ingested by the next grazing host.

The health-seeking behaviour of respondents who reported having a family member who had contracted anthrax was poor and mostly delayed, and from unorthodox sources such as traditional healers. Such delay in treatment-seeking behaviour has been reported among livestock workers, especially in other LMICs [27, 28]. It is worth noting that anthrax disease has great socioeconomic impacts on the livestock community. In this study, up to 40% were concerned that they might go out of business due to the current anthrax outbreak. Losses attributable to anthrax outbreak could be through mortality withholding of milking infected dairy herds for a period following vaccination. Other devastation may be caused by animal deaths, leading to a reduction of animal products and complete condemnation of carcasses and by-products, as well as closure of abattoirs [29].

The above findings notwithstanding, this study had some limitations. First, the performance of an on-spot assessment of the preventive practices among the livestock workers and household animal owners would have helped verify the reported claims of the respondents regarding the practice of preventive measures. Secondly, only seven of the 36 states in the country were selected for the study. However, these span the various geopolitical regions of the country. Hence the findings are generalisable and reflect the situations in other states in the various geographical regions in the country, including Niger and Lagos states, where the most recent anthrax outbreaks occurred.

## Conclusion

The study reveals a relatively high level of awareness and perception but an average knowledge level regarding anthrax among livestock workers and household animal owners in Nigeria. Significant associations were observed between the knowledge level of the respondents about anthrax and their level of education, awareness, perception as well and risk of exposure. There were important knowledge gaps, including knowledge of annual vaccination of animals, recognising common signs of anthrax in animals, and knowledge of its transmission routes. Hence, anthrax control educational programmes should target, among others, these important gaps towards achieving better outcomes. The socioeconomic impacts of anthrax expressed by the respondents in this study reiterate the need for proactive interventions from relevant stakeholders, including government, non-governmental organisations and the community.

## Data Availability

The data used in this study will be made available on request without any form of restriction. Kindly contact for any data request.

## Acknowledgments

The support of all participants who participated in the study is duly appreciated. We acknowledge the West Africa One-Health Consortium supported by the International Development Research Centre (IDRC) Canada.

## Author Contributions

C.S. conceived and designed the study. C.S., C.E., A.E.J., and A.H.K., developed the study survey tool. J.A.O., N.B.A., O.P.O., N.E., O.G.T., O.A.O., A.A.A., carried out field work and data gathering. A.E.J., I.P.M., B.M.A., A.V.O., and T.O.J carried out data analyses and visulization. A.H.K., A.E.J., C.E. conducted the quality appraisal and wrote the initial draft of the manuscript. C.S. O.S., A.R., V.C. carried out a critical review of the manuscript and all authors approved the final version.

## Notes

### Competing Interest Statement

The authors have declared no competing interest.

### Clinical Trial

Not Applicable

### Funding Statement

The author(s) received no specific funding for this work.

### Author Declarations

Ethical approval was obtained from the University of Ibadan/University College Hospital Institutional (UI/UCH) Ethics Review Committee (UI/UCH/22/0305).

